# Improving treatment precision in head and neck BNCT: delineation of oral and pharyngeal mucosa based on an MRI Atlas for standardized applications

**DOI:** 10.1101/2023.10.26.23297644

**Authors:** Katsumi Hirose, Ryohei Kato, Mariko Sato, Koji Ichise, Mitsuki Tanaka, Ichitaro Fujioka, Hideo Kawaguchi, Yoshiomi Hatayama, Masahiko Aoki, Yoshihiro Takai

## Abstract

**Background and purpose:** Boron neutron capture therapy (BNCT) has been routinely practiced for treatment of head and neck cancer in Japan. However, differences in contouring the oral and pharyngeal mucosa can lead to discrepancies in treatment. This study aimed to introduce a standardized approach using an MRI-based atlas, aiming to minimize inter-observer error and improve dose precision.

**Materials and Methods:** An MRI atlas of the head and neck mucosa was developed using water/fat-separated images from a healthy man. Using CT images from three patients, seven radiation oncologists performed contouring of the head and neck mucosa twice over a 3-week period. Contouring was first performed using CT alone, then later using fused T2-weighted images with the mucosal atlas for guidance. Contouring errors were assessed and their impacts on tumor dose were evaluated.

**Results:** The introduction of the MRI-based mucosal atlas significantly reduced inter-observer variation in mucosal volume (the coefficient of variation, abbreviated with COV, decreased from 0.61 with CT alone to 0.21 with the MRI atlas; p=0.003). Moreover, the atlas resulted in improved contour homology among observers and reduced variations in tumor dose. For all cases, COVs for maximum, mean, and minimum tumor doses were all below 5%.

**Conclusion:** Utilizing an MRI-based mucosal atlas in BNCT contouring can significantly reduce inter-observer variation, improve contour homology, and decrease variations in tumor dose. These findings suggest strong potential for standardizing and enhancing the quality of BNCT for head and neck cancer.

## Introduction

Boron neutron capture therapy (BNCT) has become a standard practice for the treatment of head and neck cancer in Japan since its approval in 2020. BNCT doses are generally prescribed based on the dose tolerable by the mucosa, which is highly sensitive to BNCT. Kankaanranta et al. adopted a physical dose of 6 Gy for the mucosal maximum dose as a dose-defining parameter in a phase II clinical trial of reactor-based BNCT for locally recurrent head and neck cancer [1]. In another phase II trial, Wang et al. indicated minimizing V_10 Gy-Eq_ of the oral mucosa as a dose constraint [2]. However, no consensus has yet been reached regarding the optimal method for contouring the oral and pharyngeal mucosa, which can significantly affect dose precision. This discrepancy potentially results in varied treatment quality across different facilities and physicians [1–4].

In previous Japanese reactor-based clinical studies, a 1-mm-thick tissue surrounding the pharyngeal air image has been generally defined as the mucosa and automatically contoured on CT images. However, this approach might underestimate mucosal contours and inadvertently result in irradiation overdose, given that the pharyngeal air-containing state varies based on the physique and posture of the patient during treatment. Such variability could compromise accurate evaluation.

The JHN001 phase I study performed during the clinical development of an accelerator-based epithermal neutron generator confirmed the safety of a maximum mucosal dose prescription of 12 Gy-Eq for BNCT in locally advanced and recurrent head and neck cancer, based on the mucosal dose that induced oral death in mice [5]. That study followed the reactor BNCT method of automatically contouring a 1-mm circumference based on the pharyngeal air-containing state. However, this method was revised in the subsequent phase II JHN002 study, in which we introduced contour extraction of the pharynx, larynx, and oral cavity mucosa as dosimetry organs using MRI signals [6]. Despite the latter method being approved in Japan, some facilities that initially performed BNCT using reactors still apply the air-containing method. Our institution argues that precise mucosa evaluation is critical for proper dosimetry within the dose tolerable by the organ, particularly since BNCT dosimetry relies on photon-equivalent doses. Hence, dose evaluations need to be based on dose tolerance reports for the mucosa of the pharynx, larynx, and oral cavity, accumulated through X-ray therapy.

This study aimed to ascertain the necessity of MRI atlas-based delineations of the mucosa in head and neck BNCT, and to scientifically verify a method for prescription dose determination that minimizes inter-observer error. Based on the scientific evidence accumulated in the above process, the goal was to disseminate and standardize this method as a treatment technique for head and neck BNCT.

## Materials and methods

We investigated whether inter-observer discrepancies in contouring the head and neck mucosa using a CT image dataset could be minimized by referring to an MRI-based anatomical mucosa atlas. Seven radiation oncologists contoured the mucosa on CT images from three head and neck cancer patients twice over a 3-week period: first using CT alone, then utilizing fused T2-weighted images and the mucosal atlas. Dose planning was conducted based on these contours to assess differences and their impacts on tumor dose. The study was approved by the institutional ethics committee.

### Preparing the MRI atlas of the head and neck mucosa

Images from MRI of a healthy man in his 70s, who had provided consent, were used to create the atlas. Images were processed using iterative decomposition of water and fat with echo asymmetry and least-squares estimation (IDEAL) on a 1.5-T Optima MR450W system (GE Healthcare Japan, Tokyo, Japan). Organ separation was achieved through water-fat separation. The IDEAL method combines asymmetrically acquired echoes and an iterative least-squares decomposition algorithm to optimize noise performance [7]. T2-weighted fast spin echo images were collected and anatomical structures were identified by comparing water images and in-phase T2-weighted images. Based on water signal intensity, the mucosal structures of the oral cavity, pharynx, and larynx were depicted on the images, and a mucosal atlas was created with reference to the literature [8, 9].

### Registration of patient image datasets

Patients were randomly selected from the treatment database of XXX Hospital after having undergone radiation therapy for locally recurrent and locally advanced head and neck cancer between 2010 and 2018. Three patients with head and neck cancer who underwent MRI within at least 3 weeks of treatment planning CT and had localized lesions with a tumor diameter of 20–40 mm confined to one side of the head and neck region were selected after consenting to the use of their treatment-related data resources.

### Delineation of head and neck mucosa by 7 radiation oncologists and dose calculation

DICOM image data were converted to 8-bit image data, the format compatible with the Simulation Environment for Radiotherapy Applications (SERA) BNCT treatment planning system, and registered in SERA [10]. A radiation oncologist specializing in BNCT delineated the tumors of the three patients mentioned above in SERA. Subsequently, seven radiation oncologists with no experience in BNCT contoured the mucosa of the same three patients in SERA. Three weeks later, the same organ was contoured on CT image data while referencing the MRI anatomy atlas, using auxiliary images fused with MRI Gadolinium T1-weighted and T2-weighted sequence data. In total, 42 sets of contour data were obtained for two conditions each in three cases of mucosal contouring by the seven observers.

### Evaluation of contour homology between observers

We defined the coefficient of variation (COV), a normalized measure of variance in the probability distribution, for each case. To assess variation in homogeneity between observers on a case-by-case basis, we defined CI*_common_* as the ratio of the common volume of all observers to the total included volume, where CI denotes the conformity index. A CI*_common_* of 1 indicates perfect agreement in volume segmentation, while a CI*_common_* of 0 indicates no overlap among the contour structures of all observers. CI*_common_* strongly depends on the number of observers, making comparison across studies challenging. Hence, CI*_gen_* was evaluated as a more universal measure [11]. This measure estimates the ratio of the sum of overlapping and non-overlapping parts between similar pairs, as follows:

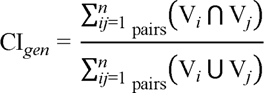

To evaluate these volume homologies, volume data were extracted from SERA log files, and CI*_common_*and CI*_gen_* were calculated using a script written in MATLAB (MathWorks, Natick, MA).

### Optimization of beam conditions and dose calculations

Dose calculations were carried out as follows. First, the beam was set up using the CT image contours of each case drawn 3 weeks prior, and photon-equivalent doses to the tumor and mucosa were calculated using a Monte Carlo code on SERA. Prescribed doses were given so that the maximum dose for the depicted mucosa was 15 Gy-eq. Calculations were performed on SERA using beam data from the NeuCure® accelerator BNCT treatment system (Sumitomo Heavy Industries, Tokyo, Japan) installed at our BNCT facility [12]. The elemental composition data set defined in ICRU report 46 was used for the transportation calculations and the derivation of absorbed doses. Relative biological effectiveness factors were set as 1, 2.4, and 2.9 for γ-rays, fast neutrons, and thermal neutrons, respectively. The compound biological effectiveness factors for ^10^B-boronophenylalanine were set as 4.9 and 4.0 for mucosa and tumor, respectively, in accordance with the JHN002 study [6]. Tumor dose was calculated using the optimized beam, considered as the CT plan. Next, using the contours obtained from the second contouring performed three weeks later, tumor dose was calculated using the same beam axis as the CT plan under beam conditions resulting in a maximum point dose to the mucosa of 15 Gy-eq. This was considered the CT/MRI atlas plan. Tumor doses in the CT plan and CT/MRI atlas plan were compared. In addition, dose differences based on the different mucosal contours contoured by seven radiation oncologists were evaluated.

### Statistical analysis

Statistical analysis was conducted using GraphPad PRISM software version 9.0 (GraphPad Software, San Diego, CA) with Student’s t-test or the Wilcoxon signed rank test, depending on data distributions. Statistical significance was defined at the level of p < 0.05.

## Results

MRI obtained using the IDEAL technique allowed mucosal structures to be clearly distinguished from other fine structures through a comparison of water images and in-phase T2-weighted images (Fig. 1A). The mucosal anatomical atlas derived from these images is shown in Figure 1B.

**Fig 1.**
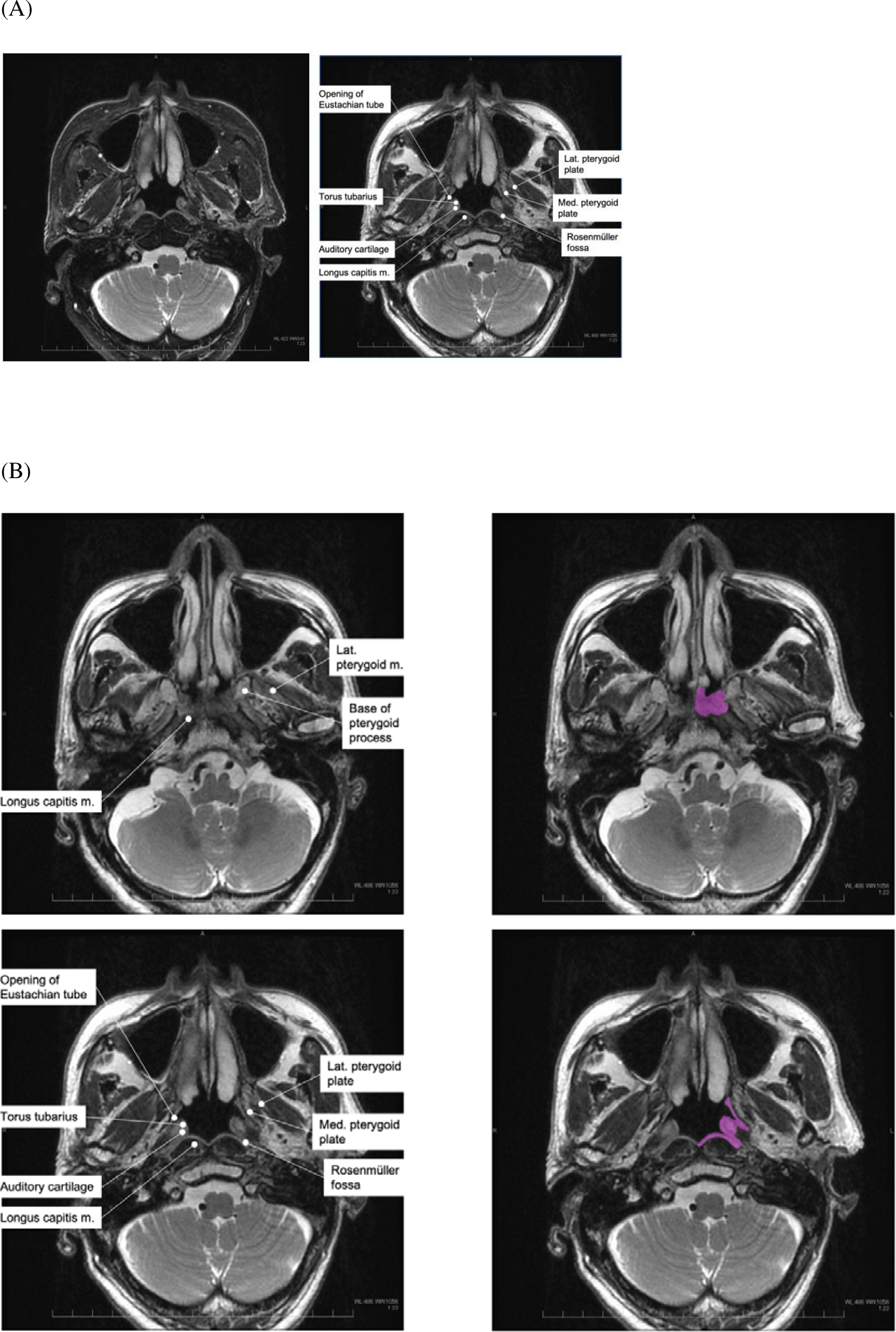
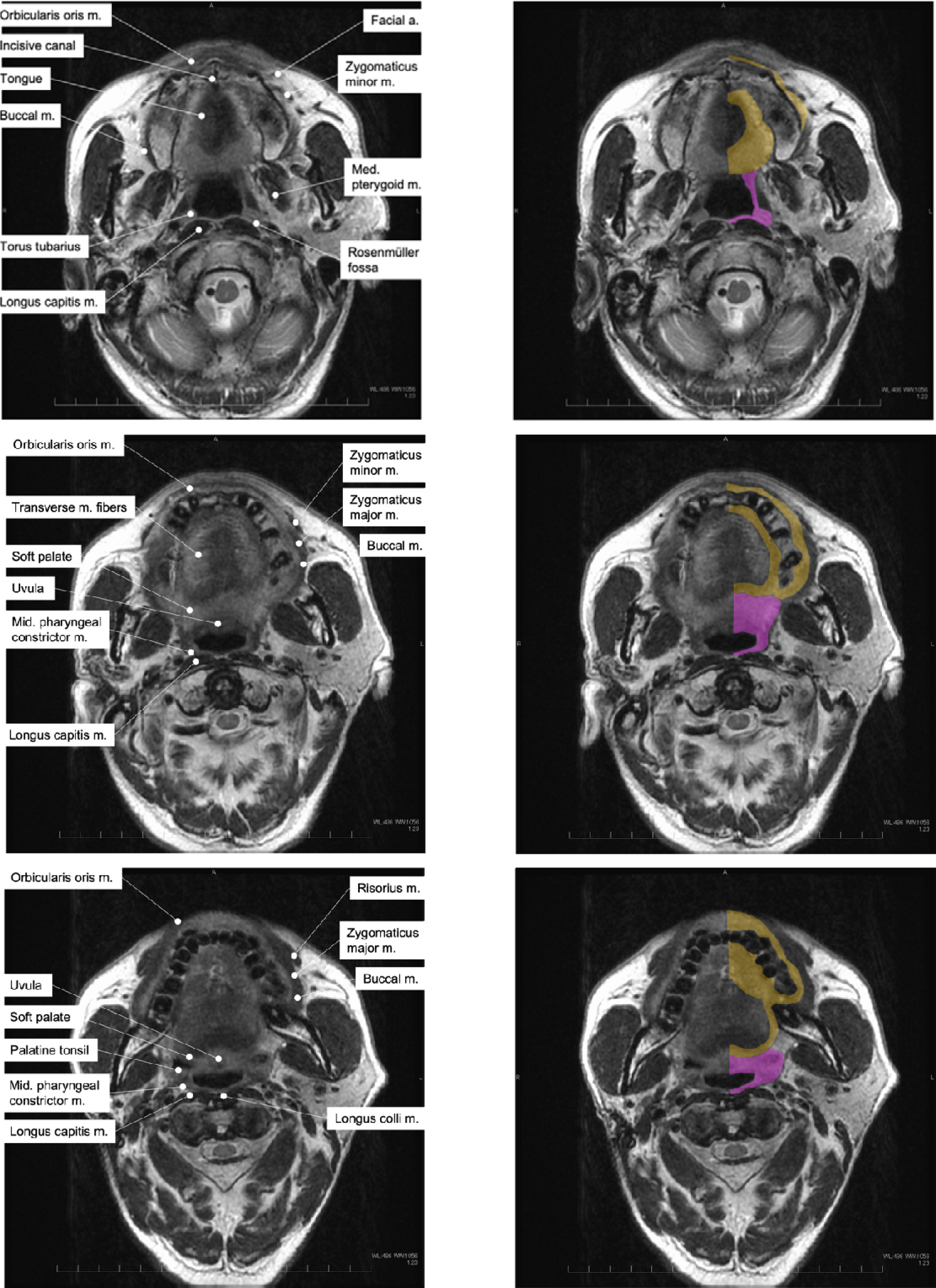
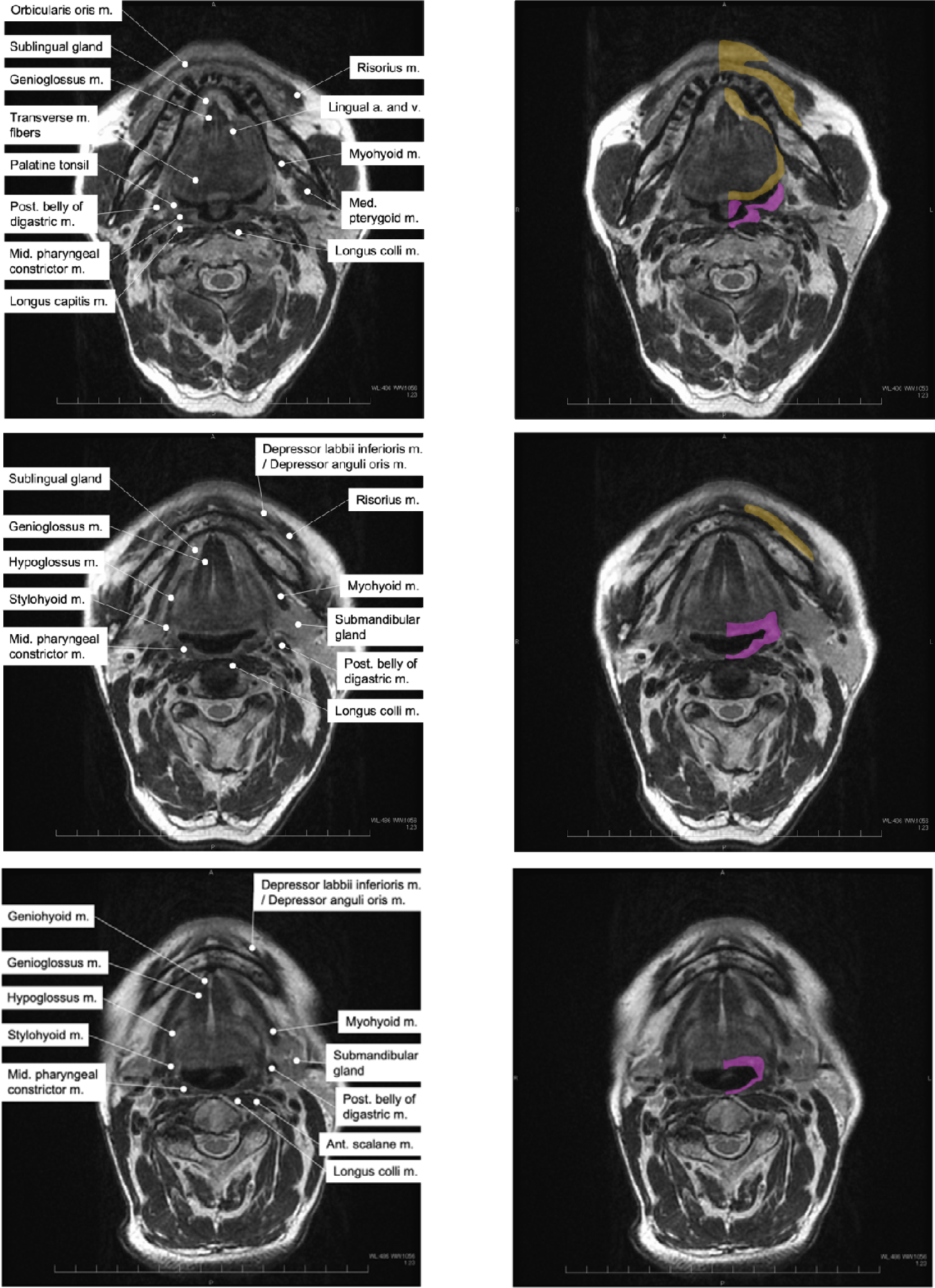
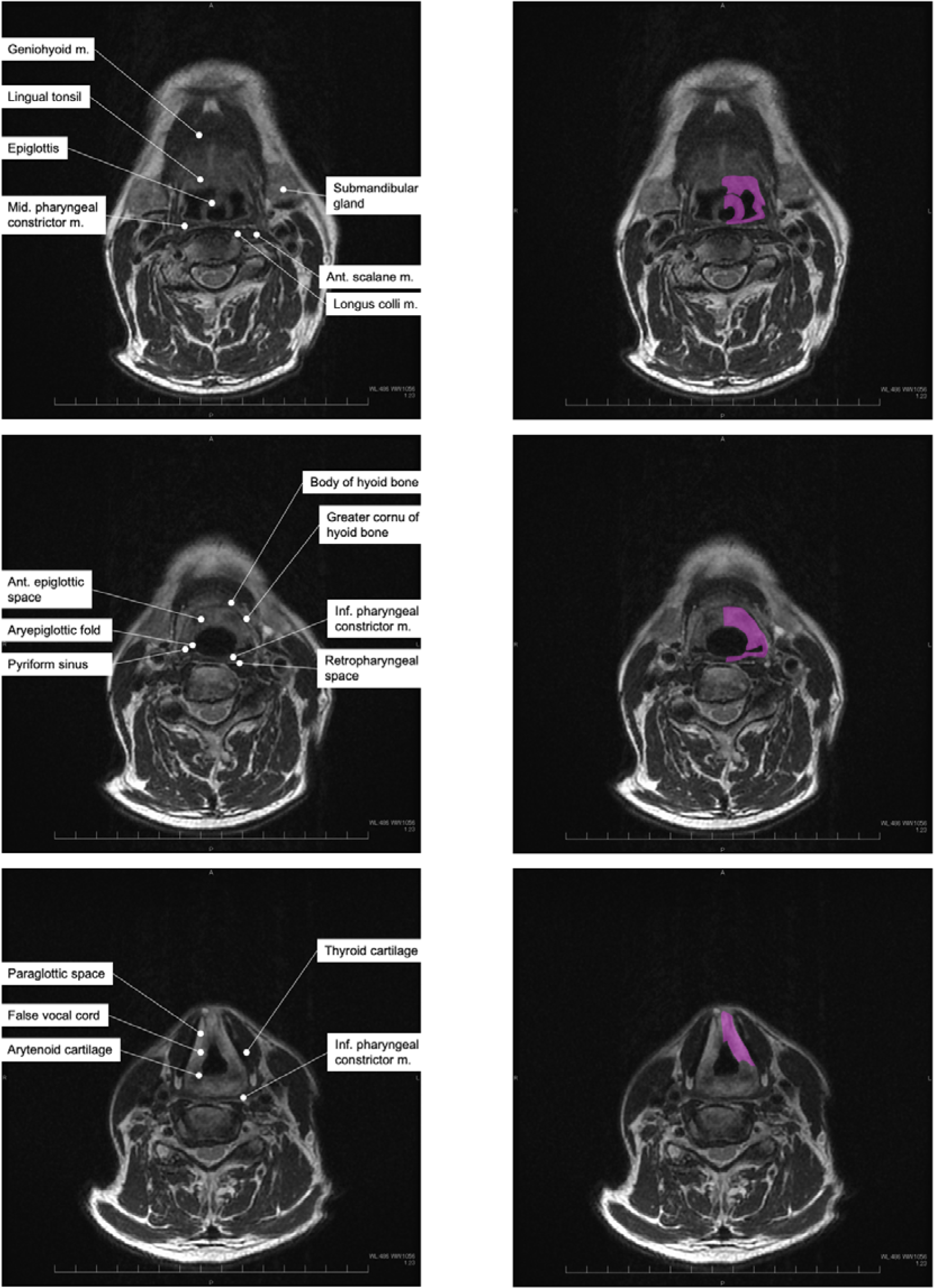
Depiction of mucosa and atlas of the head and neck mucosa A) The mucosa as depicted using the MRI IDEAL method. The left image shows uniform fat suppression for water, while the right image is an in-phase T2-weighted image. B) The atlas of the head and neck mucosa was edited by identification of the mucosa. On the right, purple and the orange outlines represent the pharyngeal and laryngeal mucosa, and the oral mucosa, respectively. The soft palate, uvula, and anterior and posterior palatal arches are outlined as oral mucosa due to the low risk of serious symptoms, such as carotid artery blowout or mediastinal abscess extension.

The patients in this study included three individuals diagnosed with locally recurrent advanced head and neck cancer, specifically hypopharyngeal cancer, parotid gland cancer, and oropharyngeal cancer. Tumor diameters ranged from 20 to 34 mm (Fig. 2A). Median cumulative clinical experience in radiation oncology for the seven radiation oncologists involved in contouring was 6.58 years (range: 2.9–29.6 years).

**Fig 2.**
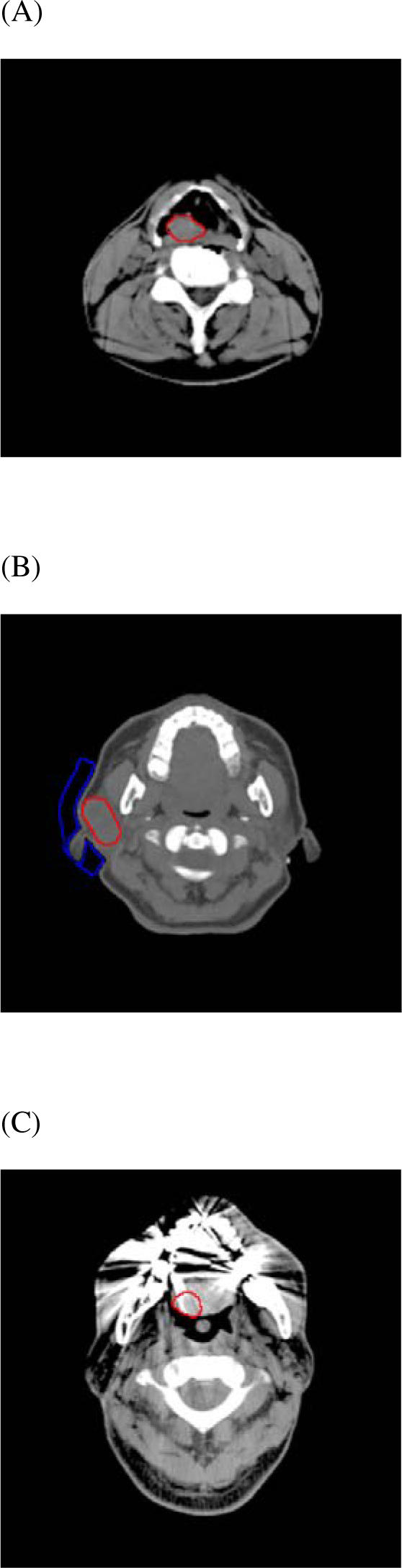
Tumor location for each case A–C) Three patients with hypopharyngeal cancer (A), parotid cancer (B), and tongue cancer (C) were included in the analysis. GTV contours in each case are shown in red.

Differences in mucosal volume among observers are presented in Figure 3. In the CT plan, each contouring observer tended to over- or underestimate mucosal volume (Fig. 3A). However, in the CT/MRI atlas plan, variations in mucosal contour volume between observers were decreased (Fig. 3B). COVs for mucosal volume in the CT plan and CT/MRI atlas plan were 0.61 ± 0.02 and 0.21 ± 0.05, respectively, being significantly lower in the CT/MRI atlas plan (p = 0.003, Fig. 3C). Mucosal contours by the seven observers for patient 2 are shown in Figure 4. CI*_common_* was 0.02 ± 0.01 in the CT plan and 0.12 ± 0.04 in the CT/MRI atlas plan. On the other hand, CI*_gen_* was 0.258 ± 0.02 in the CT plan and 0.47 ± 0.05 in the CT/MRI atlas plan. The CT/MRI atlas plan exhibited reduced variations in shape among observers and improved homogeneity, although some variability remained, reflecting the complexity of the mucosa (Fig. 4B).

**Fig 3.**
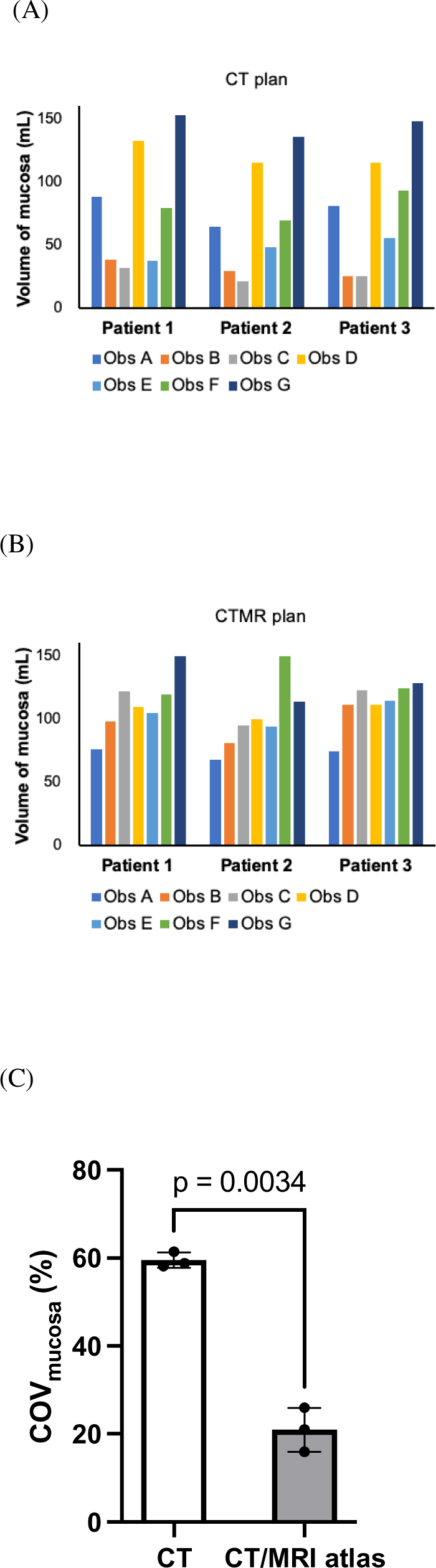
Differences in mucosal volume between observers Mucosal volumes in the CT plan (A) and CT/MRI atlas plan (B) were compared among observers. C) The COVs of mucosal volume were compared between the CT and CT-MRI plans.

**Fig 4.**
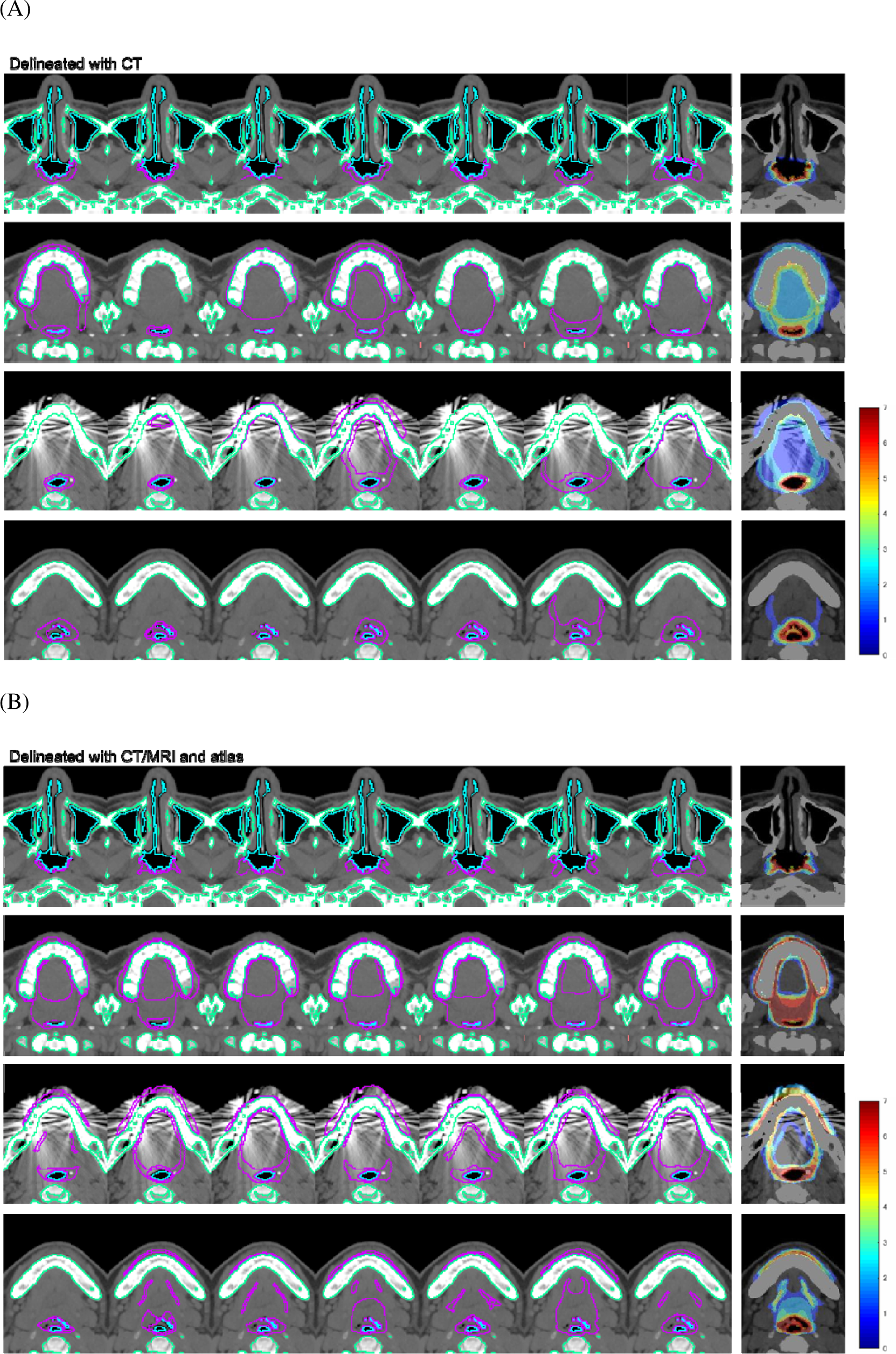
Overlap of mucosal contours between observers for Case 3 The degree of overlap of mucosal contours was depicted using different color tones in the CT plan (A) and CT/MRI atlas plan (B). The rightmost image shows the degree of mucosal overlap between observers. From the leftmost image to the right, mucosal contours by Observers A–G were lined up.

For BNCT dose calculations, the beam condition of each case was determined from typical positions of entry and exit points on the skin and distances from the skin surface to the beam aperture used in actual clinical practice in our BNCT facility (Fig. 5A). Regarding tumor dose, the CT plan showed significant variation in tumor dose for Cases 2 and 3 compared to Case 1 (Fig. 5B, C; Table 1). This was due to the variable contouring of the oral cavity and the proximity of the parotid tumor (Case 2) or tumor at the tongue root (Case 3) to the oral cavity, causing large variations in the prescribed tumor dose (Fig. 4). On the other hand, Case 3 with hypopharyngeal tumor located in the lower neck, away from the oral cavity, showed less dose difference between observers. In the CT/MRI atlas plan, variations in tumor dose were reduced, and COVs for tumor maximum, mean, and minimum doses were below 5% in all cases (Table 1). That is, the results suggest that the CT-MRI plan dramatically improved the magnitude of variability in each case, and inter-observer error for tumor dose was suppressed to within a level acceptable for radiotherapy.

**Fig 5.**
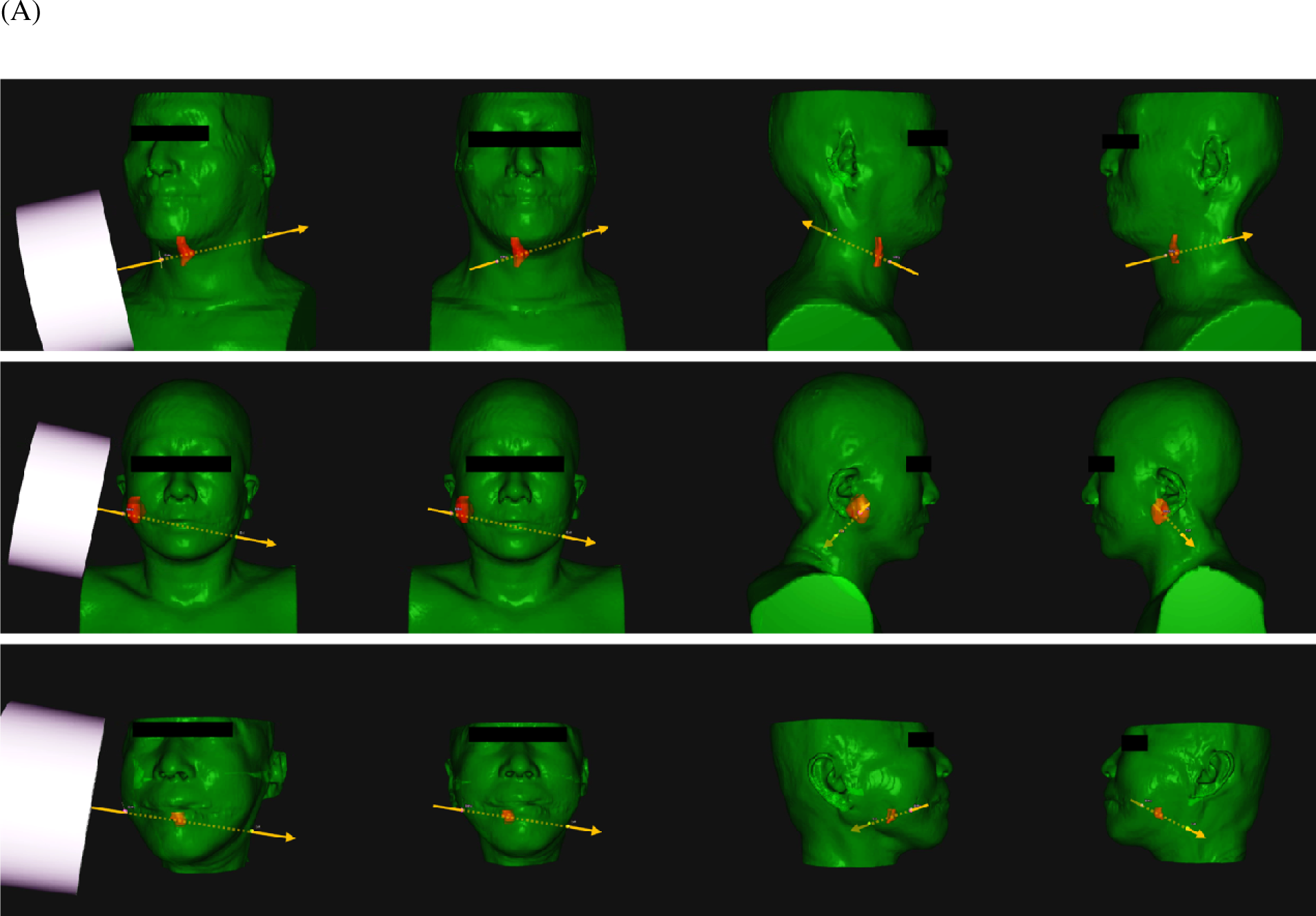
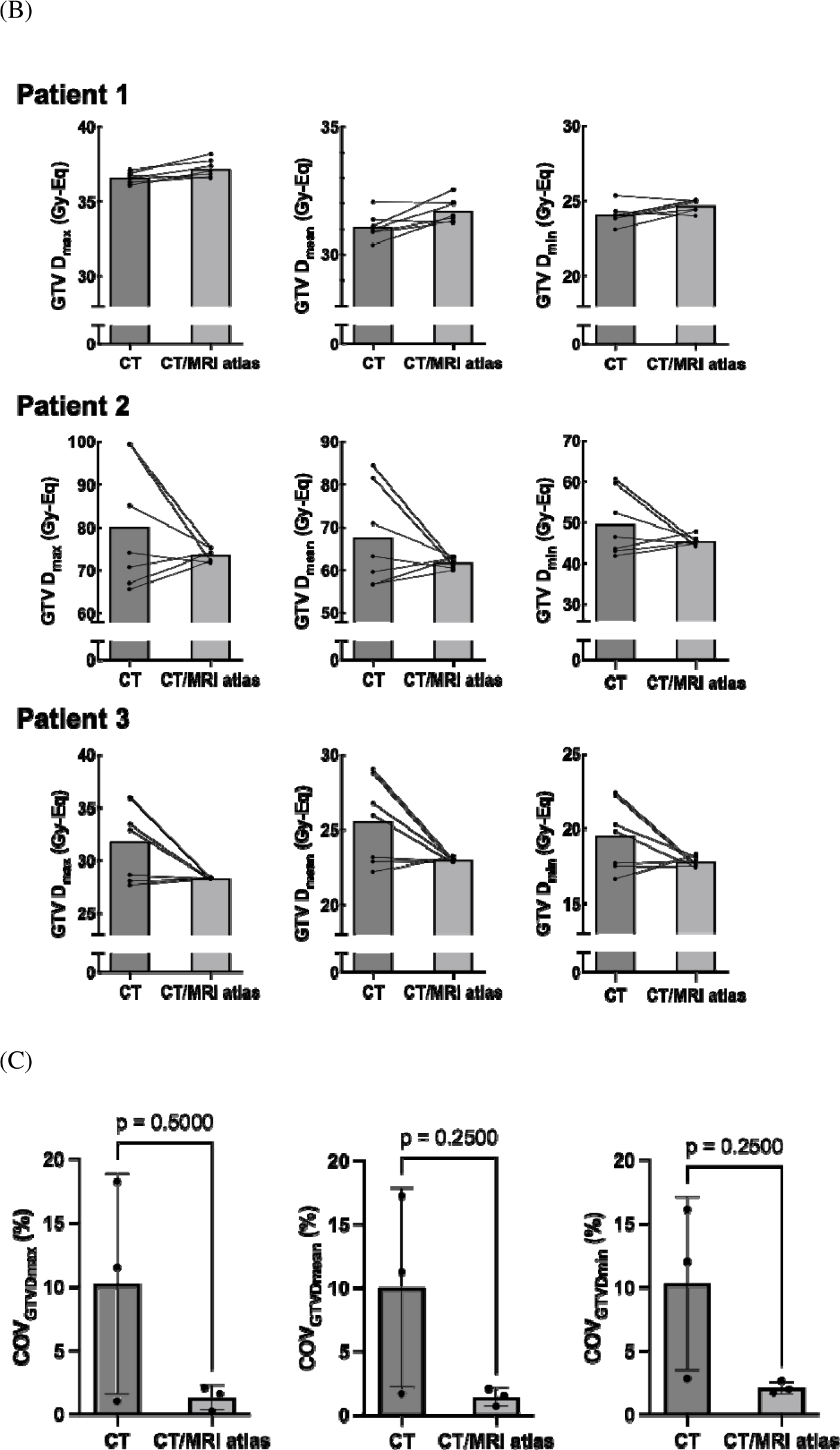
Beam condition and inter-observer variation in tumor dose for each case A) Body surface contours and GTV as visualized using RayStation®. Axes are depicted passing through the entry and exit points of neutron beam centers on the body surface. Images in the upper, middle, and lower rows are for the cases of hypopharyngeal cancer, parotid cancer, and tongue root cancer, respectively. B) Comparison of GTV D_max_, D_mean_, and D_min_ between the CT plan and CT/MRI atlas plan. C) Comparison of COVs for the GTV D_max_, D_mean_, and D_min_ between the CT plan and CT/MRI atlas plan.

**Table 1.** Improvement of inter-observer variation for tumor dose by mucosa delineation with the MRI atlas.

| COV | CT | CT/MRI atlas |
| --- | --- | --- |
| *Patient 1* |  |  |
| GTV D<sub>min</sub> | 2.82 | 1.68 |
| GTV D<sub>mean</sub> | 1.67 | 1.53 |
| GTV D<sub>max</sub> | 1.02 | 1.64 |
| *Patient 2* |  |  |
| GTV D<sub>min</sub> | 16.13 | 2.67 |
| GTV D<sub>mean</sub> | 17.21 | 2.09 |
| GTV D<sub>max</sub> | 18.23 | 2.11 |
| *Patient 3* |  |  |
| GTV D<sub>min</sub> | 12.04 | 1.95 |
| GTV D<sub>mean</sub> | 11.23 | 0.68 |
| GTV D<sub>max</sub> | 11.52 | 0.25 |

Abbreviation: COV, coefficient of variation; GTV, gross tumor volume; D<sub>min</sub>, minimum dose; D<sub>mean</sub>, mean dose;
D<sub>max</sub>, maximum dose.

## Discussion

In IMRT for head and neck cancer, mucosal dose is deemed a risk factor for stomatitis [13, 14]. Nonetheless, oral mucosa contouring is ambiguously described, and the pharyngeal mucosa is often neglected in target evaluations [15]. As severe pharyngeal mucositis may lead to eating disorders, dysphagia, and tube feeding, inclusion of the pharyngeal mucosa in evaluations is crucial. This study demonstrated that a mucosal anatomy atlas could reduce observer discrepancies in contouring the pharyngeal region.

Although usage of the MRI atlas reduced mucosal contouring errors, some variability persisted, as demonstrated by CI values. However, significant improvements were obtained in the uniformity of the tumor dose parameter. Considering the relationship between contour shape and mucosal prescription points, a similar shape of oral and pharyngeal mucosal structures is sufficient for evaluation. The CI for mucosal contours using the MRI atlas was at the same level as the degree of CI for prostate contours delineated using cone-beam computed tomography [17]. The mucosa is a complex structure and this improvement comparable to that for the CI of the prostate, a much simpler structure, signifies the benefits of the MRI atlas.

The interpretation of mucosal shape in the oral cavity varies considerably among physicians, resulting in completely different dose-prescribed BNCT implementations when treating tumors near the oral cavity without guidelines or certain regulations. This study noted that in the case involving the hypopharynx, which is more distant from the oral cavity, differences in tumor minimum dose between physicians were maintained within a COV of 3% in CT-based delineation. Conversely, in cases of parotid and tongue cancers, differences in tumor dose prescription among physicians reached up to 16%, since irradiation was performed in the vicinity of the oral mucosa. An extremely high tumor dose was given to the parotid gland tumor by a physician who neglected the mucosa of the oral cavity. When the CT plan is approved for treatment, the mucosa dose depicted using the MRI atlas, which is considered relatively close to the “truth”, was 19.8 Gy-eq, substantially exceeding the original 15 Gy-eq. This extremely high dose to the mucosa of the oral cavity will carry a high risk of severe mucositis and mucosal defects. Past BNCT, performed with the mucosa contoured as a 1-mm thickness around the oral cavity, might have led to cases of mucosal irradiation overdose. In contrast, the JHN002 study, using MRI-based mucosal contouring as in this study, proved very safe, with only one case of grade 3 mucositis, and the frequency and grade of stomatitis remained lower than in any other report [6]. Therefore, strictly defining the method for delineating the mucosa is crucial for future BNCT clinical trials. Currently, a safety cohort study for dose optimization of accelerator-based BPA-BNCT in patients with unresectable locally recurrent squamous cell carcinoma of the head and neck (the ST-BNCT2001 study; ClinicalTrial.gov identifier no. NCT05883007) using the MRI mucosa atlas is ongoing.

Properly delineating complex mucosa structures is time-consuming. This study initially planned to examine at least five cases, but the time-consuming nature of delineating mucosal contours resulted in a focus on only three cases. Each CT plan took 23 ± 12 min and each CT/MRI plan took 34 ± 7 min to delineate the mucosa, with each radiation oncologist spending 172 ± 48 min per case. This limitation led to only three cases being analyzed in this study. Despite this limitation, the data strongly suggest that using an MRI atlas could considerably improve dosimetric variations in lesions near the oral cavity.

In conclusion, using an MRI atlas to delineate the regions of oral, pharyngeal, and laryngeal mucosa resulted in decreased inter-observer variation and increased consistency. Consequently, COVs for tumor dose were reduced. These findings suggest the utility of a mucosal anatomy atlas for BNCT dose planning in head and neck cancer. Further research outcomes using this method are expected to enhance the efficacy and safety of BNCT for head and neck cancer.

## Data Availability

All data produced in the present study are available upon reasonable request to the authors

## Acknowledgements

This study was supported by a Grant-in-aid for Research on Radiation Oncology from the Japanese Society for Radiation Oncology, 2018–2019.

## Notes

### Competing Interest Statement

Katsumi Hirose declare the personal research grant from Stella pharma corporation. All other co-authors declare no conflict of interest.

### Funding Statement

This study did not receive any funding

### Author Declarations

Ethics committee of Southern Tohoku Hospital gave ethical approval for this work

